# A simple, cost-effective and extraction-free molecular diagnostic test for sickle cell disease in noninvasive buccal swab specimen for a limited-resource setting

**DOI:** 10.1101/2022.06.14.22276141

**Authors:** Priya Thakur, Pragya Gupta, Nupur Bhargava, Rajat Soni, Narendra Verma, Sangam Giri Goswami, Gaurav Kharya, Vinodh Saravanakumar, Padma G, Suman Jain, Jasmita Dass, Mukul Aggarwal, Sivaprakash Ramalingam

## Abstract

Sickle cell disease (SCD) is the most prevalent life threatening blood monogenic disorder. Currently, there is no cure available, apart from bone marrow transplantation. Early and efficient diagnosis of SCD is key to disease management which would make considerable strides in alleviating morbidity and reducing mortality. However, the cost and complexity of diagnostic procedures like Sanger sequencing impede the early detection of SCD in a resource-limited setting. To address this, the current study demonstrates a simple and efficient proof-of-concept assay for detection of patients and carriers from extraction free non-invasive buccal swab samples by isothermal <u>D</u>NA <u>A</u>mplification coupled <u>R</u>estrictase-mediated cleavage (iDAR). This study is a first of its kind reporting the use of buccal swab specimens for iDA in molecular diagnosis of a genetic disease all the while being cost effective and time saving with the total assay time of around 150 minutes at a cost of USD 5. Further, iDAR demonstrates 91.5% sensitivity and 100% specificity for detecting all three alleles; SS, AS and AA having 100% concordance with Sanger sequencing. The applicability of iDAR assay is further demonstrated with its adaptation to a one-pot reaction format, which simplifies the assay system. Overall, iDAR is a simple, cost-effective, precise and non-invasive assay for SCD screening with the potential for use in a limited resource setting.

## 1. Introduction

Sickle cell disease is a monogenic hematological disorder with varying clinical phenotypes. SCD is caused by a point mutation in the beta globin *(HBB)* gene found on chromosome 11. This single base pair substitution causes aberrant hemoglobin (HbS), which leads to the characteristic sickle form of the red blood cells (RBCs).

SCD is exceedingly prevalent in sub-Saharan Africa, Southeast Asia and the Middle East. According to the World Health Organization (WHO), approximately 300,000 infants were born, with SCD in these countries. ^1,2^ Without treatment, many patients born with SCD die before the age of five.^3^ Early and efficient diagnosis of SCD would enable better management and alleviate morbidity and reduce mortality. Molecular based screening specifying the genotype of SCD is crucial not only for diagnosing patients but also for genetic counseling to reduce the spread of SCD.

The cost and complexity of current standard diagnostic procedures impede the early detection of SCD in resource-limited settings across the world. Currently, methods such as isoelectric focusing (IEF), electrophoresis and high-performance liquid chromatography (HPLC) are used to detect the wildtype (AA), sickle cell carrier (AS) and sickle cell disease (SS) genotypes. The above tests are expensive and require highly trained staff to interpret the results.^4^ Moreover, HPLC cannot stand alone as a diagnostic test and must be done along with a confirmatory test such as sequencing based DNA analysis before a final diagnosis can be made.^4,5^ Though different point-of-care assays are available commercially, most are antibody tests that cannot be applied in individuals who have undergone blood transfusions in the last 90 days, as it may result in misleading interpretation due to presence of hemoglobin proteins from the donor blood. Antibody tests also prove inconclusive in cases where newborns undergo blood transfusion due to multiple reasons including hemorrhagic shock, and premature anemia, among other birth related complications,^6^ requiring re-validation 90 days later further delaying diagnosis and increasing cost. In addition, all newborns express basal level fetal hemoglobin in the first several months of their life. As a result, in order to establish a definite diagnosis, the levels of the different forms of hemoglobin must be compared at a later period.^7,8^

Blood samples have long been utilized in clinical diagnosis and genetic investigations including for newborn screening. However, blood samples are susceptible to clotting, smearing and oversaturation.^9^ Hence genetic screening using blood samples needs to be collected very carefully and has to be done by trained professionals. In addition, blood collection is intrusive, especially for pediatric patients, and it may not be essential for molecular diagnosis, where easy-to-collect, less-expensive alternatives such as buccal swabs and saliva can be attempted.

Allele-specific identification of disease mutation can provide classifications of genetic disease for precise diagnosis. Traditional techniques to identify point mutations are mainly dependent on methods such as gold standard DNA sequencing^10^ and real-time PCR, ^11^ which are often time-consuming and need costly instruments and a clean environment. Isothermal DNA amplification approaches have similar sensitivity and specificity as a conventional PCR, and can be set up at constant temperature and requires only a less expensive heating block.

Thus far, the majority of the PCR-based genetic tests relies on the use of the purified genomic DNA from clinical samples,^12^ which constitutes a major bottleneck in molecular diagnosis. In the current pilot study, a unique strategy is demonstrated that allows specific discrimination of point mutation using iDAR. A simple non-invasive buccal swab specimen has been adopted, which is economical and can be used as a direct sample matrix without DNA extraction process. Moreover, the assay has been simplified to a one-pot single tube reaction of a complete sample-to-answer workflow which could facilitate the genetic analysis of the clinical sample in less than 150 minutes. For a proof-of-concept, the robustness of iDAR assay in detection of allele-specific SCD genotypes from extraction-free non-invasive crude lysates of buccal swab samples has been shown with patient, carriers and wild type samples.

## 2. Materials and methods

### 2.1 Construction of recombinant HBB plasmids carrying HBB wild-type (AA), and sickle cell mutation (SS) sequence

The HBB gene was amplified from buccal swab samples of a healthy individual/wild type (AA), and SCD patients (SS) using HBB primers (HBBF: CTAGGGTTGGCCAATCTACTC and HBB-R: AGTAATGTACTAGGCAGACTGTG). The PCR amplicons were cloned into pJET plasmid (Fermentas, USA) following the manufacturer’s instructions. Subsequently, the ligated products were transformed into *Escherichia coli* DH5α and colonies were screened by colony PCR. Recombinant plasmids were isolated and purified using Plasmid MiniPrep Kit (Thermo Fisher Scientific, USA). It was further confirmed through Sanger sequencing. The recombinant HBB plasmids were termed pSR-β^AA^ and pSR-β^SS^ and the concentration of plasmid DNA were estimated using NanoDrop 2000 (Thermo Fisher Scientific, USA).

### 2.2 Design of iDA primers

iDA primers for the target gene HBB were designed as per manufacturer’s recommendation (TwistDx, UK). The primers were designed based on following parameters as iDA product size between 250 to 300 bp, primer size between 30 to 35 nucleotides, and primer GC content 40 - 70% using IDT OligoAnalyser tool. The iDA primers designed were HBB iDA forward primer: GTCAGGGCAGAGCCATCTATTGCTTACATT, HBB iDA reverse primer: ATAGACCAATAGGCAGAGAGAGTCAGTGCC. All the primers used in this study were synthesized from Sigma-Aldrich, India.

### 2.3 Sensitivity and specificity analysis

The HBB gene was amplified using HBB-F and HBB-R primer and the concentration of the amplicon was determined using the Nanodrop 2000 (Thermo Fisher Scientific, USA). The copy number of the amplified product was calculated using the formula: Number of copies per µL = ((Amount of ds DNA) X (6.022×10^23^ molecules per mole)) / ((length of dsDNA) × 10^9^ ng X (660 g/mole)). The PCR amplified product was serially diluted to obtain the copy number per µL in a range of 10^8^-10^0^. Each dilution was used for iDA. Simultaneously, real time quantitative PCR (qPCR) was also performed for each dilution using the Premix ExTaq Master Mix (Takara Bio Inc. Japan) according to the manufacturer’s protocol.

### 2.4 Buccal swab samples collection

The study was conducted after obtaining approval from the Institutional Ethics Committee at the Council of Scientific and Industrial Research -Institute of Genomics and Integrative Biology (CSIR-IGIB), New Delhi and All India Institute of Medical Sciences (AIIMS, New Delhi). Written informed consent was obtained for the study. Participants were requested to give samples using nylon-flocked buccal swabs (Himedia, INDIA) to swab each inner cheek at least 10-15 times. Buccal swab samples were collected and immediately placed in the lysis buffer.

### 2.5 Processing of buccal swab specimens in different lysis buffers

Four different lysis buffers namely Buffer A (custom-made buffer), buffer B,^13^ buffer C (custom-made buffer) and buffer D, ^14^ were attempted for the direct amplification of clinical buccal swab samples. The details of buffer compositions are given in Table 2. Buccal swab samples collected in lysis buffer A, B and C were directly used for performing iDA. However, for lysis buffer D, the samples were vortexed for 10 minutes and then incubated at 95°C for 10 minutes keeping buccal swabs in the tube itself. Samples were allowed to cool down at room temperature and buccal swabs were removed from the tube. Finally, 120 µL 1M Tris-Cl (pH-8.0) was added. If the iDA was not performed immediately, samples were stored at 4°C. The buccal swab samples which were collected and transported as dry swabs were processed in lysis buffer D on arrival and further processed in the similar manner as fresh buccal swabs.

### 2.6 iDA assay setup and electrophoresis

iDA reactions were performed using the TwistAmp Basic kit (TwistDx Limited, UK, TABAS03KIT) according to the manufacturer’s instructions. For a 50 µL reaction, 37.5 µL of master mix containing 29.5 µL of rehydration buffer, 2.4 µL of 10 µM HBB iDA forward primer, 2.4 µL of 10 µM HBB iDA reverse primer (all together called HBB iDA mix), 5-10 ng of recombinant plasmid or 5-20% of buccal swab lysate, was sequentially added to a reagent pellet and the solution was pipetted up and down 6 to 8 times to mix. Subsequently, 2.4 µL of magnesium acetate (MgOAc) was added to the lid, the tubes were closed and mixed by brief vortexing. After brief centrifugation, the reaction was immediately initiated by incubating in a heat block with a constant temperature (either 42°C or 37°C) for 20 minutes. The iDA products were analyzed on 2% agarose gel electrophoresis and documented using a UV transilluminator or gel documentation system.

### 2.7 Targeted cleavage of iDA products

Eco81I-mediated cleavage of iDA products were performed in 30 µL reaction volume containing 4-8 µL of direct iDA product, 1X Tango buffer, and 15U of Eco81I (Thermo Scientific, USA). Alternatively, DdeI enzyme can also be used for iDAR. The reaction was incubated at 37°C for 1 hour, and the cleaved products were analyzed on 2.5% agarose gel electrophoresis. To ensure the purity of the SCD iDA product, Nco1 enzyme (New England BioLabs, USA) was used as an internal digestion control.

### 2.8 PCR amplification and Sanger sequencing

Clinical buccal swab samples were used for HBB gene amplification using primer HBB-F (seq): GCTGTCATCACTTAGACCTCAC and HBB-R (seq): CATAGACTCACCCTGAAGTTCTCA that covers SCD mutation using standard PCR with OneTaq DNA polymerase (NEB, USA). The PCR products were column purified using a PCR purification kit (Genetix Biotech India) and were subjected to Sanger sequencing. Sequencing was carried out at AgriGenome Labs Pvt. Ltd., Kochi, India

### 2.9 One-pot-reaction

For the one-pot, one reaction of HBB iDA mix was divided into 5 reactions, each containing 8 µL of HBB iDA mix and 5-20% of buccal swab sample. The reaction was activated by addition of 0.8 µL MgoAc and subsequently amplified at 37°C for 10 minutes and inactivated at 85°C for 5 minutes in the heat block. The digestion premix for the 30 µL reaction was made with 1X Tango buffer, 15U Eco81I, and added to the respective tubes for mutation screening. As an internal control, a digestion premix containing 1X Cutsmart buffer and 15U of NcoI was prepared for 30 µL reaction. One-pot products were analyzed on 2.5% agarose gel electrophoresis and documented using a UV transilluminator or gel documentation system..

## 3. Results

### 3.1 iDAR strategy

The experimental strategy used for the development of a simple and efficient allele-specific genetic diagnostic test for detecting SCD mutation in non-invasive buccal swab samples is shown in Figure 1A. Clinical samples were collected in the form of buccal swabs and treated with a lysis buffer to release the nucleic acids followed by iDA. SCD mutation in the HBB gene eliminates the restriction enzyme Eco81I site. The Eco81I restriction enzyme cleaves the amplified product in the sequence CCTNAGG (where N represents any nucleotide). Hence, when thymine (T) is replaced with adenine (A), it alters the target recognition site for Eco81I. There are three possible outcomes of Eco81I cleavage depending on the genotype of the individual (AA, AS, SS) and the expected cleavage patterns are depicted in Figure 1B. The banding patterns formed by the enzyme cleavage after separation indicate the genotype as follows. (i) In wild-type (AA) healthy individuals with AA, the amplicon is cleaved by Eco81I and produces two bands. (ii) In sickle cell heterozygous (carrier) individuals with AS, no cleavage is expected in the S allele, however, the A allele gets cleaved, resulting in three bands. (iii) In SCD patients SS, there is no cleavage due to the mutation in both alleles, so a single parent band appears in the gel. In order to ensure the purity of SCD iDA products, an internal control, Nco1 enzyme was used in the study which will cleave the iDA product irrespective of their genotype and generate two bands. Genomic structure of HBB gene and the sequence to which iDA primer binding sites are indicated by arrows is shown in Supplemental Figure S1

**Figure 1.**
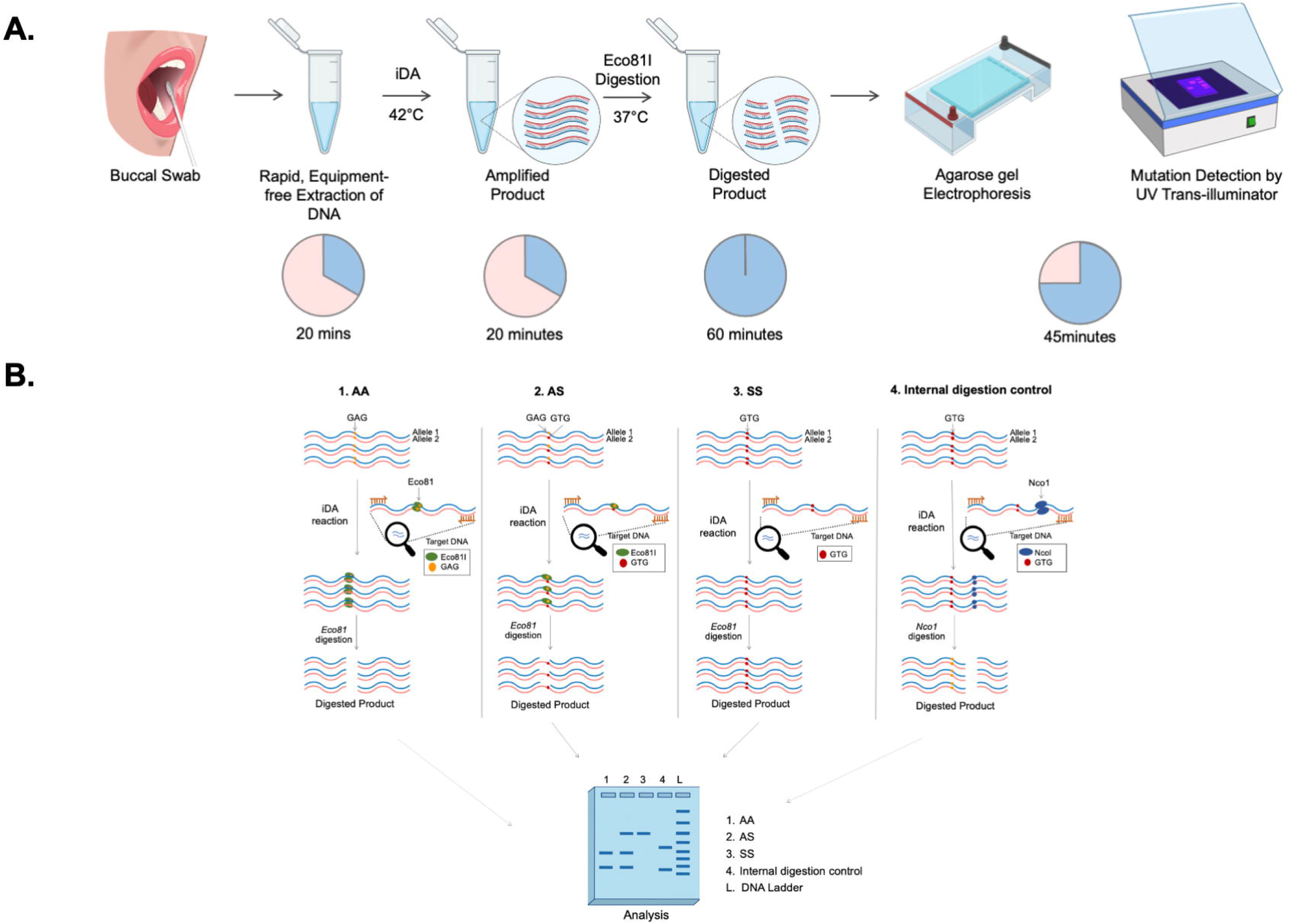
Experimental design of iDAR assay. A. Schematics showing the workflow of iDAR. B. Schematic showing the screening strategy for allele specific sickle cell mutation detection using iDAR.

### 3.2 Optimization of iDA with different buccal swabs lysates

First, iDA based amplification was examined using recombinant plasmid, pSR-β^AA^ and pSR-β^SS^ mimicking the genotypes of AA, and SS in 10 µL reactions at 37°C, 40°C and 42°C. The results demonstrated amplification at all the temperatures with equal intensity at expected size (Figure 2A). In order to reduce the processing steps, crude buccal swab lysates were employed for the study. The release of nucleic acid is very important for iDA hence use of an appropriate lysis buffer would be crucial for consistent and efficient iDA using buccal swabs. In this study, previously reported lysis buffers and custom-made lysis buffers were compared for iDA using buccal swabs collected from healthy study participants (AA). All 4 lysates that were used in the study were processed and added to iDA reactions and the results suggested that iDA performed with Lysis buffer D was most sensitive consistently (Supplemental Figure S2, Table 1).

**Figure 2.**
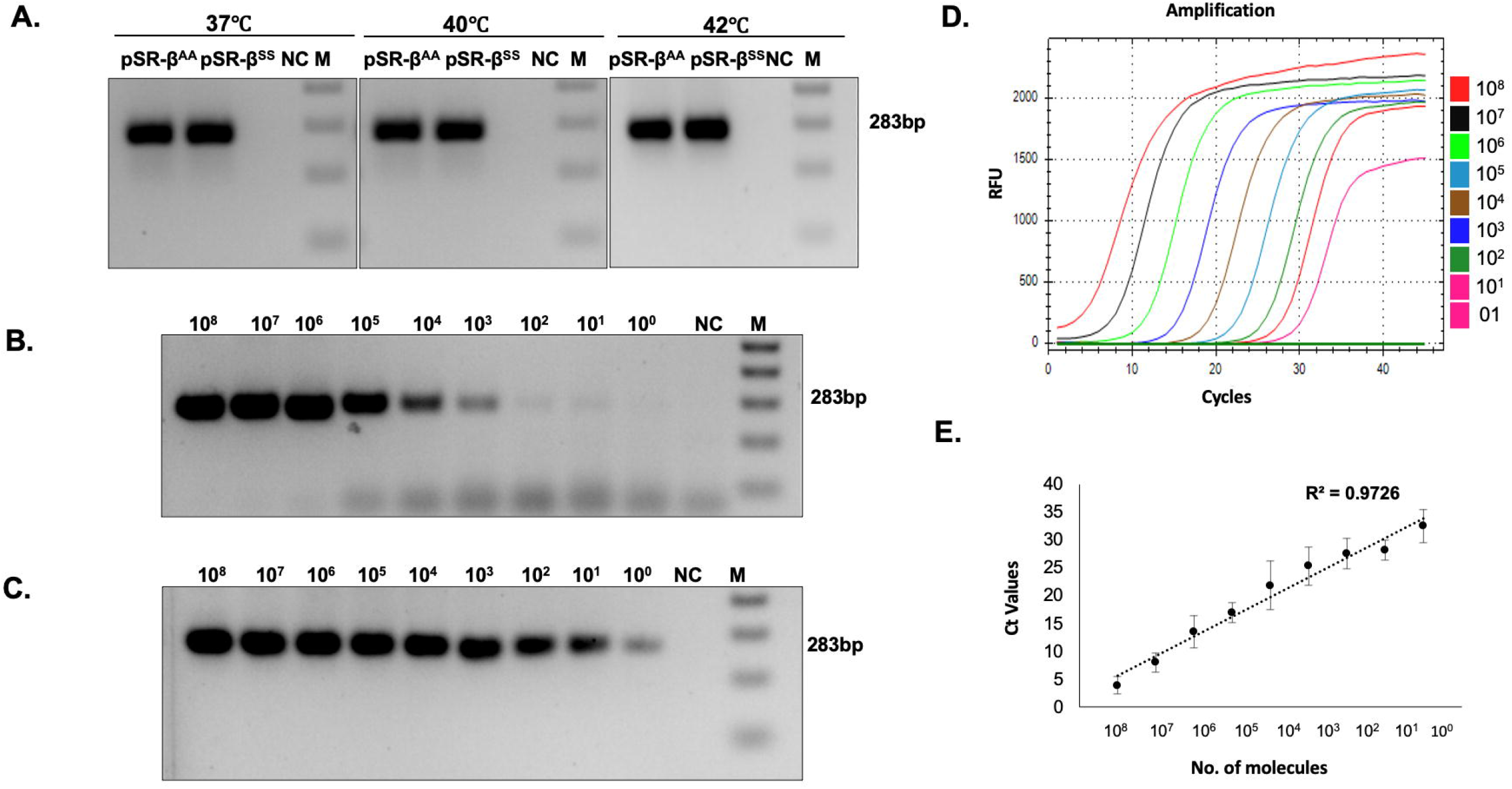
Optimization of iDA using recombinant plasmids. A. iDA of recombinant plasmids pSR-β^AA^ (WT) and β^SS^ (MT) at different temperatures. Minimum copy number required for (B)iDA, (C, D E) qRT-PCR for n=3

**Table 1.**
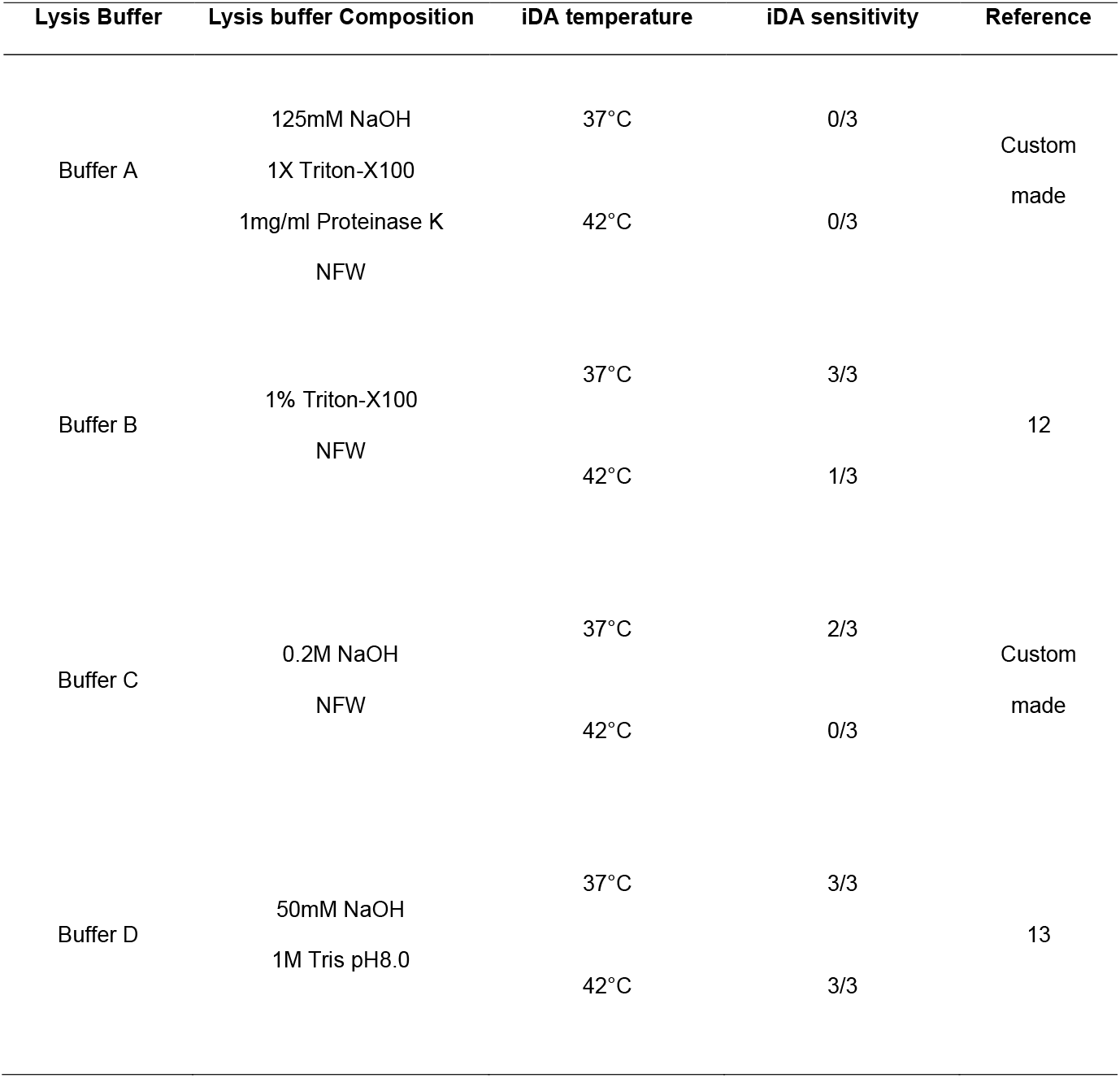
A comparative analysis of different lysis buffers for iDA.

### 3.3 Limit of detection

In the present study, the limit of detection was assessed and compared with currently available qRT PCR. A range of 10^8^-10^0^ copies/ul dilution was used to demonstrate the sensitivity of the assay. The results suggested that iDA is capable of amplifying from as low as 10-100 target DNA copies (Figure 2B). In comparison, qRT-PCR being a more sensitive and sophisticated technique was able to detect 1-10 DNA copies (Figure 2C, D and E), however, it is relatively more expensive and requires trained professionals. Overall, the above results suggest that iDA is a relatively sensitive technique that can be used as an alternative of the pre-existing sophisticated molecular diagnostic tools in low-resource settings.

### 3.4 Effect of temperature, time and concentration of buccal swabs lysates on iDA

To enhance the efficiency of target amplification, different parameters such as (i) the effect of temperature (ii) the reaction time and (iii) the inhibitory effect of the buccal swab lysates on the reaction efficiency was taken into account. First, the inhibitory effect of direct buccal swab lysate on the iDA reaction efficiency was examined in the range from 5% to 30% of buccal swab lysates in the total reaction volume. The results demonstrated successful target amplification at all concentrations of buccal swab lysate (Figure 3A and B). However, beyond a point, the concentration of buccal swab lysate in total reaction volume inhibited the amplification. Thus, it is suggested that 5%-20% of direct buccal swabs lysate is optimal for the iDA. We also compared the amplification sensitivity of buccal swab lysate between more sensitive qRT PCR and iDA. A Low (2%) and high (20%) buccal swab lysate concentration of 5 clinical samples with unknown copy numbers were used and the results demonstrated that iDA worked in all samples at both lysate concentrations (Figure 3C), however, in qRT PCR few samples at higher percentage of buccal swabs lysate inhibited the amplification (Figure 3D).

**Figure 3.**
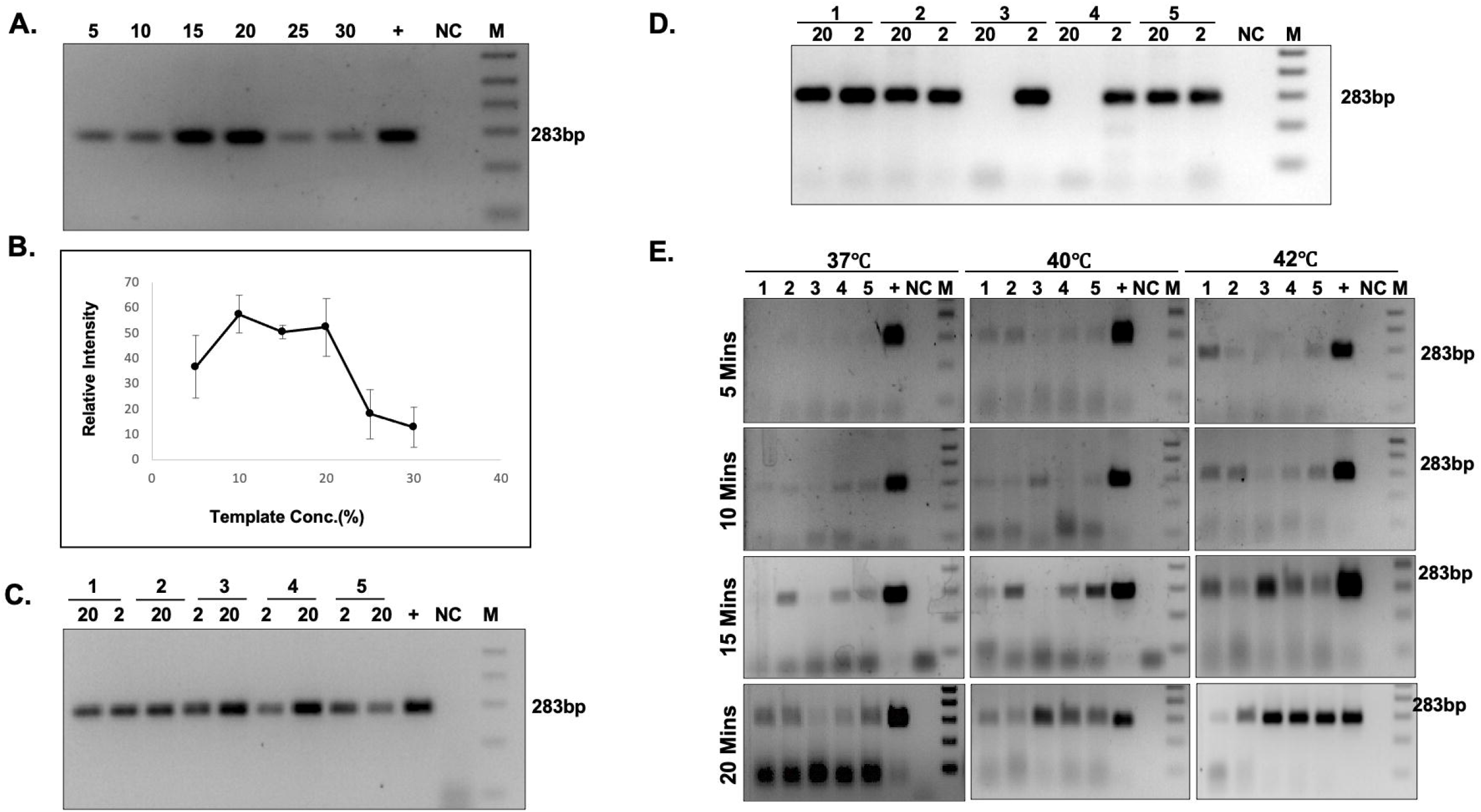
Effect of temperature, time and concentration of buccal swab lysates on iDA. A. iDA with varying buccal swab lysate concentrations (%) and (B) relative quantification=3. Amplification sensitivity with high (20%) and low (2%) buccal swab lysate concentration in (C)iDA and (D) qRT-PCR; n=5 (E) Sensitivity of iDA at different temperatures and timepoints n=5.

Next, the effect of temperature and time on the amplification efficiency was assessed. The iDA was performed at 37°C, 40°C and 42°C for 10 minutes, 15 minutes and 20 minutes each. The results demonstrated that target DNA amplification at all three temperatures and reaction times using iDA (Figure 3E, Table 2). However, enhanced amplification of the target DNA was consistently observed at 42°C with reaction time of 20 minutes.

**Table 2.** Effect of amplification time and temperature on iDA

| iDA Temperature | iDA Amplification Time | iDA sensitivity |
| --- | --- | --- |
| 37°C | 5 mins | 0/5 |
|  | 10 mins | 0/5 |
|  | 15 mins | 3/5 |
|  | 20 mins | 4/5 |
| 40°C | 5 mins | 0/5 |
|  | 10 mins | 1/5 |
|  | 15 mins | 4/5 |
|  | 20 mins | 5/5 |
| 42°C | 5 mins | 1/5 |
|  | 10 mins | 4/5 |
|  | 15 mins | 5/5 |
|  | 20 mins | 5/5 |

In order to bring down the assay cost, we hypothesized that reducing the iDA per-test reaction volume would lower the assay cost significantly. We investigated reaction volumes ranging from 10 to 50 µL ul and the appropriate concentration of the buccal swab lysate was added to the reaction and found that amplification was observed in all quantities tested in the above range without impairing iDA performance (data not shown).

### 3.5 Successful iDAR using clinical buccal swab samples

Detection of the genotype (AA, AS and SS) analysis is based on the cleavage pattern observed upon Eco81I digestion. Hence, iDA was coupled with restrictase (iDAR) to ultimately deduce the clinical status of the study participants. In this study, first recombinant plasmids PSR-β^AA^ and pSR-β^SS^ were used to validate iDAR. The results showed complete cleavage in AA and no digestion in SS, as expected (Figure 4A). We then subjected iDA from pSR-β^SS^ plasmid for Nco1 digestion to verify the quality of iDA product. As expected, Nco1 digestion resulted in complete cleavage (Figure 4A).

**Figure 4.**
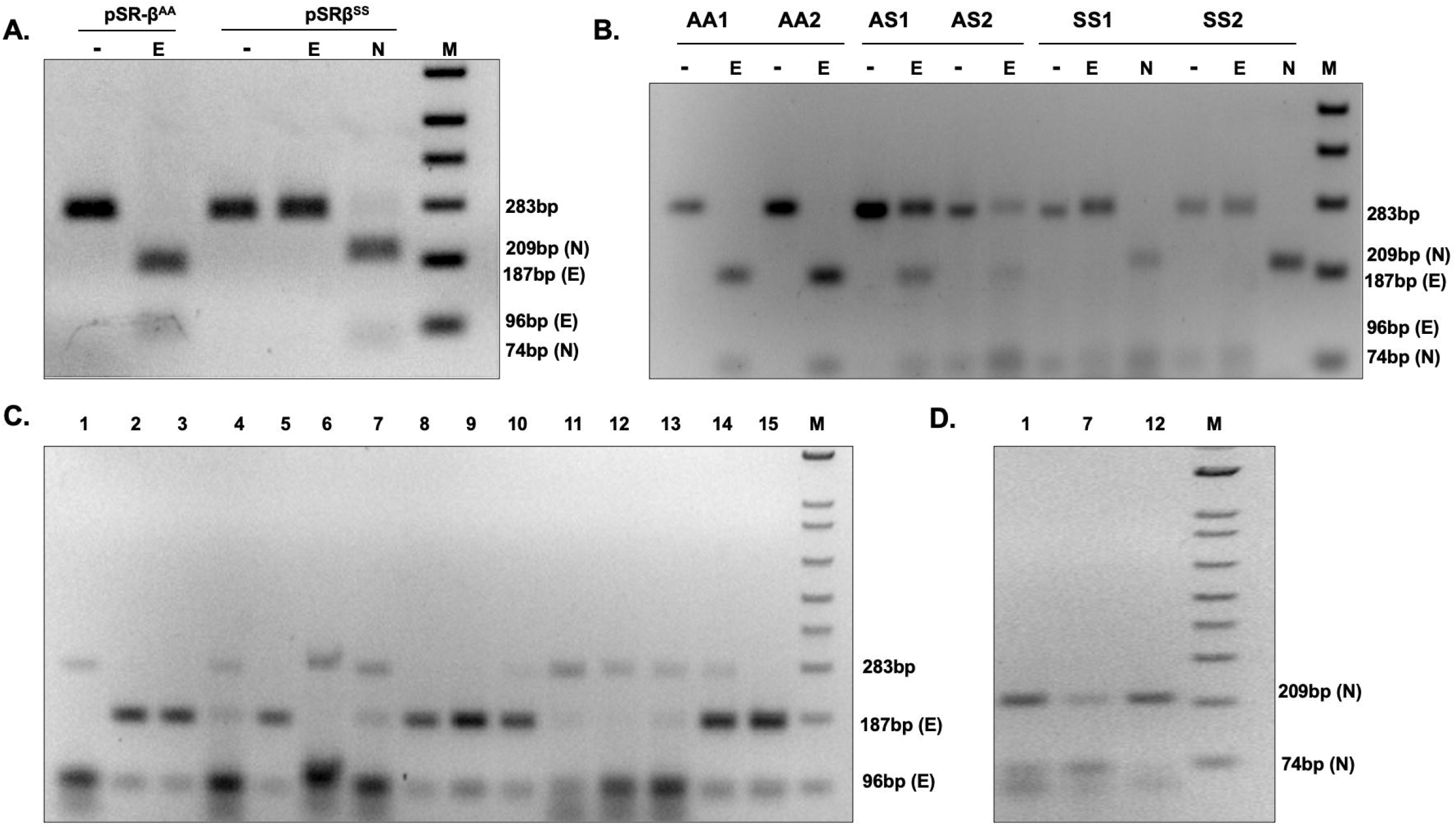
iDAR using recombinant plasmids and patient buccal swabs: A. iDAR using Eco81I (E) digestion of recombinant plasmids pSR-β^AA^ and pSR-β^SS^ with internal digestion control of NcoI (N) B. Eco81I screening in individuals with known genotypes AA, AS^S^ and SS^S^) with internal digestion control of NcoI (N) for n=2. C. iDAR of buccal swabs of unknown genotype; n=15. D. iDAR with internal control digestion with Nco1(N) of SS samples

Next, to verify the reliability of the optimized iDAR assay, we further evaluated using clinical samples. The buccal swab samples were collected with lysis buffer D and subjected to direct iDA amplification at 42°C for 20 minutes. Subsequently, 4-8 µL of iDA product was directly subjected to Eco81I digestion at 37°C for 1 hour. However, FastDigest Eco81I requires only 15-30 minutes for digestion. The major advantage of this iDAR assay is, it provides the qualitative answer in terms of “yes” or “no”. Representative amplification and cleavage from clinical samples of three genotypes, AA, AS and SS with two samples each are shown in Figure 4B. Finally, 15 randomly selected samples from the above clinical pool were further evaluated using Sanger sequencing. Sequencing results corroborated with the iDAR, which proved the accuracy and specificity of our assay. Evaluation of our iDAR assay using 100 clinical samples showed 91.5% sensitivity and 100 % specificity (Table 3). The sensitivity of the assay was increased, by modulating the concentration of the buccal swab sample in total reaction volume. By doing so, we were able to achieve the sensitivity of the assay to almost 100% (data not shown). Storage of the buccal swab lysates at 4°C for a prolonged period of time before processing did not affect iDA sensitivity. However, it was observed that multiple rounds of processing of the same lysate results in reduced iDA sensitivity. Further, a blind iDAR experiment was performed with 15 randomly selected buccal swab lysates from the above pool (Figure 4C). Furthermore, Nco1 digestion was carried out on Eco81I digestion negative samples to conclude SS genotype (Figure 4D). Genotype deduced based on Eco81I and Nco1 cleavage pattern corroborated with the known genotype of the study participants.

**Table 3.**
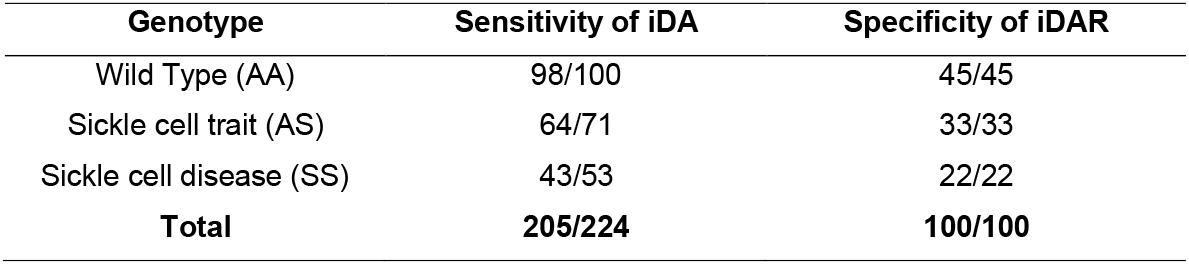
Sensitivity and specificity of the iDAR assay.

| Genotype | Sensitivity of iDA | Specificity of iDAR |
| --- | --- | --- |
| Wild Type (AA) | 98/100 | 45/45 |
| Sickle cell trait (AS) | 64/71 | 33/33 |
| Sickle cell disease (SS) | 43/53 | 22/22 |
| **Total** | **205/224** | **100/100** |

We also evaluated our iDAR assay system with buccal swab samples stored under dry conditions at different temperatures and processed at different time points as the study aims to provide confirmatory genetic diagnosis in sickle cell carrier and SCD patients in low- and mid-income countries. It is advantageous as dry swabs can be collected in non-clinical settings, making the transportation and handling of the samples easy for people living in remote areas without any requirement of cold storage. In order to assess the effect of dry swabs in prolonged storage, 5 dry swab specimens from each study participant were collected and stored at room temperature for 4 different time points 12 hr, 24hr, 48 hr and 72 hr. The iDA with stored dry swab samples produced consistent amplification at all four -time points (Supplemental Figure S3).

Next, we checked the impact of storage temperature of buccal swabs samples on iDA sensitivity. This was done as countries like India experience very high temperatures which may impact sample integrity. For this, we collected samples from 5 study participants and stored them at 25°C, 37°C and 42°C for 5 days. iDA was performed for these samples and our result suggests successful amplification for all study participants at all above temperatures. (Supplemental Figure 4, Table 4).

**Table 4.** Effect of long-term incubation of buccal swabs at variable temperatures on iDA sensitivity.

| Storage Temperature | Number of days of storage | iDA sensitivity |
| --- | --- | --- |
| 25°C | 5 | 5/5 |
| 37°C | 5 | 5/5 |
| 42°C | 5 | 5/5 |

### 3.6 Successful demonstration of One-pot reaction

In this study, the genotype analysis of clinical samples using direct buccal swab specimens has been shown till now. However, independent iDA followed by restrictase screening separately may result in cross contamination, if multiple samples are handled together avoids any contamination and to ease the workflow, we hypothesized if iDAR could be performed in the same reaction tube (One-pot reaction) (Figure 5A.). Since, our previous results have shown successful amplification at 37°C for both recombinant plasmids and clinical samples (Figure 1A and Figure 3E), we first checked if iDAR can be optimized by reducing the reaction volume to 5 uL and amplification time to 10 mins, followed by 60 mins restrictase screening using recombinant plasmids, pSR-β^AA^ and pSR-β^SS^ at 37°C in one-pot reaction. Results showed expected Eco81I cleavage pattern for AA and SS with visible intensity. Further, Nco1 internal control digestion also demonstrated expected cleavage pattern for SS ensuring the quality of iDA. (Figure 5B). we tested one pot reaction with clinical buccal swab lysates of known genotype following the above experimental strategy. Initially, 5 uL of HBB iDA mix was used with clinical buccal swab lysates, however, results showed inconclusive digestion pattern due to less target DNA amplification (data not shown). Hence, one-pot reaction was tested with 10uL of HBB iDA mix that resulted in expected Eco81I cleavage pattern with visible intensity followed by NcoI digestion of SCD clinical samples (Figure 5C) showing the effectiveness of One-pot iDAR for the diagnosis of clinical samples

**Figure 5.**
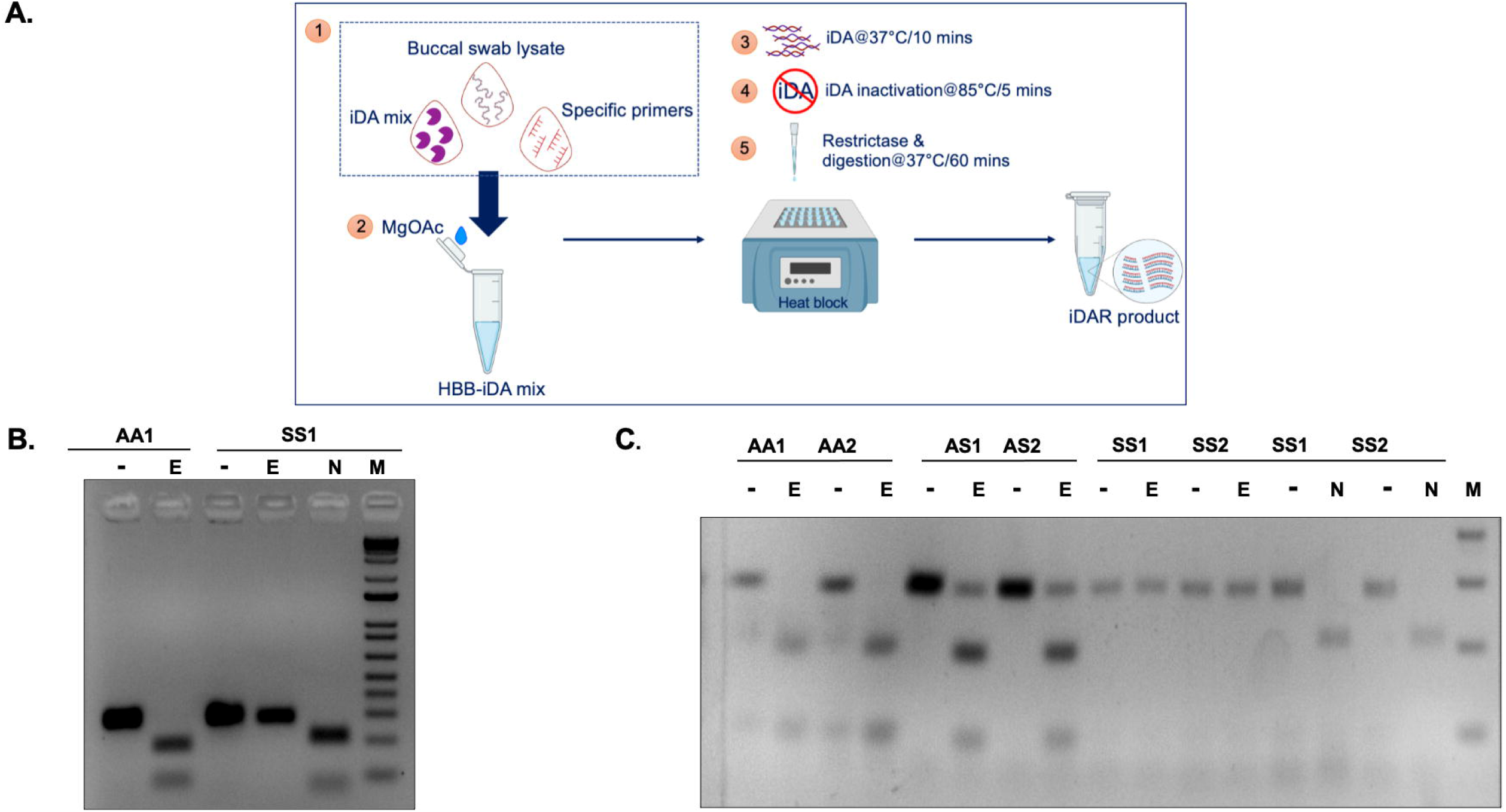
One-pot iDAR A. Schematics showing steps in One-pot reaction. B One-pot iDAR using recombinant plasmids pSR-β^AA^ (AA) and SS (SS). C One-pot iDAR using Eco81I (E)screening in individuals with known clinical genotypes (AA, AS and SS) with internal digestion control of NcoI (N) for n=2.

## 4. Discussion

Sickle cell disease has emerged as a significant global health burden and to combat it, early diagnosis with genetic confirmation is pertinent. One way of doing it is by enabling simple and accurate diagnostic screening in a limited resource setting leading to early diagnosis, expeditious management in affected newborns as well as aid in identification of carriers and subsequent genetic counseling.^15^

Currently, in order to make a conclusive diagnosis there are a series of tests that are done, a full blood panel followed by HPLC and further confirmation of the genetic mutation through Sanger sequencing. Although the latter ensures an accurate diagnosis, it is not quick and easily deployable, requiring specialized instruments and trained professionals. Hence developing simple and efficient strategies of diagnosis is crucial.

SCD screening can also be achieved by several point-of-care tests, such as assays based on hemoglobin solubility ^16^ and monoclonal antibody-based detection of hemoglobin AA or SS, ^17^ which are becoming accessible to reduce the necessary infrastructure challenges and costs. However, any assay based on monoclonal antibodies such as HemoTypeSC,^17^ that identifies hemoglobin proteins cannot be utilized in patients who have just been transfused recently, since blood transfusions contain globin proteins from the donor and it leads to misinterpretation of the results.^4^ Solubility testing cannot differentiate between SCD and carrier, and hence is not recommended due to these limitations and is less ideal for SCD screening.^16^

In recent years, lateral flow assays (LFAs) have broadly been used as portable molecular diagnostic methods. In 2015, Kanter et al developed a Sickle SCAN based on lateral flow immunoassay to detect SCD.^8^ However, the assay has several limitations, such as misinterpretation due to visual reading, polyclonal antibody cross-reactivity, inconsistency in the band’s intensity, and false-positive results in identifying HbAS with HbSS. Hence, the authors suggested further confirmatory validation. Another study by Natoli et al have reported a strategy for allele-specific detection of sickle cell mutation using blood samples.^18^In their assay, two separate reactions for each sample are required to detect AA and SS genotypes, which is the major limitation of the above study.

Isothermal DNA Amplification based molecular diagnosis is an emerging strategy that has been successfully employed for screening of infectious diseases such as Ebola virus disease (EVD) and COVID-19.^19,20^ Current study focuses on developing an isothermal DNA amplification-based strategy for screening of SCD. There are however certain challenges that had to be addressed in order to develop this assay such as the nature of sample collected and quickest way that can be processed without compromising the efficiency and accuracy of the test.

Our iDAR assay system is very simple and easily deployable in a resource limited setting, as it only requires a heat block and a tube prefilled with a mixture containing the essential reagents for amplification and subsequently treated with restrictase in the same tube for screening. Notably, addition of direct buccal swab samples does not significantly affect the sensitivity of the iDA. It was noted that the sensitivity of iDA was compromised when more than 20% of the buccal swab lysates make up the reaction volume. Present study has demonstrated the sensitivity of iDA to be 91.5%, however the sensitivity of the assay can be increased by changing the concentration of buccal swab lysate in the reaction volume. Moreover, we were able to achieve 100% amplification by modulating the buccal swab lysate concentration. In addition, we also observed that iDA assay detected the genotype correctly even in the patients, who had undergone blood transfusion within a few weeks before sample collection.

In this study, a single test reaction is required to differentiate all the three genotypes; SS, AS and AA alleles, which brings down the assay cost. By leveraging Eco81I-mediated sequence-specific cleavage, our iDAR assay system shows a more consistent and precise outcome for heterozygous (AS) and homozygous (SS) samples. The iDAR assay offers an alternative to DNA sequencing or real-time PCR for rapid SCD genotyping while keeping the advantages of simplicity, affordability and a faster qualitative readout.

Traditionally, blood samples have been used for clinical diagnostics and genetic studies. However, blood collection is an invasive procedure, both expensive and needing trained professionals. Even though blood is the preferred sample for running most of the molecular tests, using it for DNA amplification is not suitable for multiple reasons.^21,22^ The nucleic acid isolated from blood or blood containing samples is usually low and therefore the sensitivity of isothermal amplification reduces. A more suitable and less invasive alternative to blood is taking saliva for use in molecular diagnosis. However, certain study participants have difficulty in spitting into a tube, while others struggle to produce enough saliva for genetic testing ^23^ Dry mouth is a typical medication side effect that can make saliva collection difficult.^23^ However, buccal swab sample collection is quick, non-invasive, and inexpensive, taking all this into account this study sought to explore the feasibility of performing SCD genotyping. It would also make it easy for patients and their family members to collect samples at clinics or at their homes. Biological samples such as stool, serum, plasma, vaginal, and nasal have been used for isothermal amplification, ^24^ however, this is the first report where buccal swab specimens have directly been used for iDA especially in the molecular diagnosis of a genetic disease.

Developing a molecular diagnostic test that does not require DNA isolation is difficult since existing genotyping strategies based on DNA amplification necessitate relatively high DNA purity. ^25,26^ Hence, developing a genetic test that analyses the samples without DNA isolation and purification has remained elusive. The current study aimed to establish an inexpensive low-complexity buffer system with routine laboratory chemicals that make direct iDA feasible by utilizing a simple procedure that allows for the efficient release of DNA from buccal swab specimens.

Previously, to aid the release of nucleic acids from human cells, various procedures have been applied, such as using complex solvents and expensive unique materials. ^27,28^ In the iDAR assay system, we leveraged NaOH to lyse the cell membrane for releasing DNA and Tris-Cl as a crucial pH control agent and provided an appropriate environment for iDA reaction. Thus, the iDAR assay system proposed in this study is capable of genotyping target genes without undergoing the laborious, time-consuming and expensive nucleic acid isolation and purification step, avoiding the possibility of cross contamination.

Till date, no information is available on the stability of DNA in “dry swab” for storage and transport at various temperatures and its impact specifically on the iDA for genetic testing. The storage of biological samples is a major challenge during sample collection at a remote place, as sample integrity is a factor of great importance. Ideally biological samples must be kept in temperature-controlled, adequately qualified storage ^29^ and be processed as soon as possible after collection. But in a realistic scenario, the vast majority are stored and transported some distances before processing. In this study, the viability of dry swabs without any transport reagent was tested for storage and transport at various incubation times and temperatures. It was observed that dry swabs can be transported maximum at 42°C and upto 72 hours without affecting iDA. These findings are significant for low- and middle-income nations that have limited access to rapid refrigerated transport and storage of samples, which is a cost-effective option. Finally, this study reveals the practical and financial benefits of employing dry swabs without transportation reagent.

In the iDAR assay system, reagents account for the majority of the overall per-test cost. Thus, it was hypothesized that lowering the per-test reaction volume can significantly reduce the assay’s cost. Thus, different reaction volumes were experimented and it was observed that amplification with direct buccal swab specimens as templates in all volumes worked efficiently without compromising assay sensitivity and performance. Hence, it brings down the amplification cost by one fifth and also significantly lowers the overall assay cost to USD 5.

Although iDAR is able to detect all the three genotypes; AA, AS and SS, it cannot discriminate among AA, Hemoglobin Type C genotypes AC and CC (HbC), thus, future work on exploring the development of an integrated assay system would be beneficial. Another limitation is the differentiation of SS from Sβ thalassemia which is recognized as heterozygous AS genotype.

In summary, the iDAR assay platform has considerable advantages. The results show that iDAR is an efficient assay capable of detecting all three genotypes SS, AS and AA. Existing genetic diagnostic tests based on PCR such as DNA sequencing, and qPCR, typically require a complicated operational procedure, as well as expensive and sophisticated instruments that may not be available in many laboratories, whereas the iDAR assay can be done with less complex instruments and provides reliable, inexpensive, easy-to-use and affordable genetic diagnostics for SCD and facilitate the expansion of SCD screening in low-resource settings.

## Data Availability

All data produced in the present work are contained in the manuscript

## Acknowledgements

This work was supported by the Council for Scientific and Industrial Research (CSIR) grants (MLP2010 and HCP023) and Department of Biotechnology grant (BT/GET/119/SP31652/2020) Government of India (GoI) to SR. PT, PG, SG and NB are recipients of Senior Research Fellowship from Department of Biotechnology, Council of Scientific and Industrial Research, University Grant Commission and Indian Council for Medical Research, GoI respectively.

