## Supplementary figures and images for "A simple, cost-effective and extraction-free molecular diagnostic test for sickle cell disease in noninvasive buccal swab specimen for a limited-resource setting"

### SuppSlide1.tiff

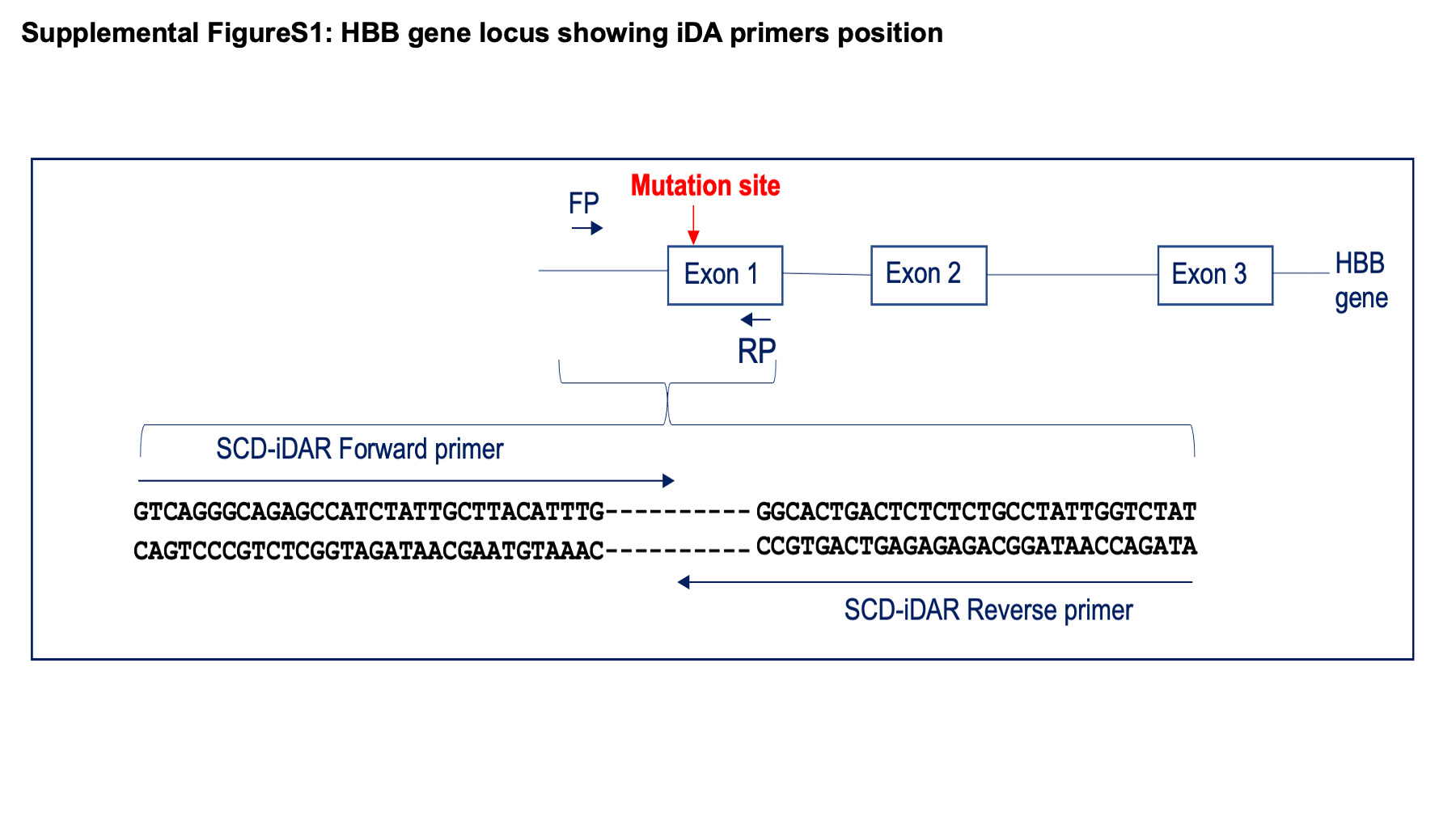

### SuppSlide2.tiff

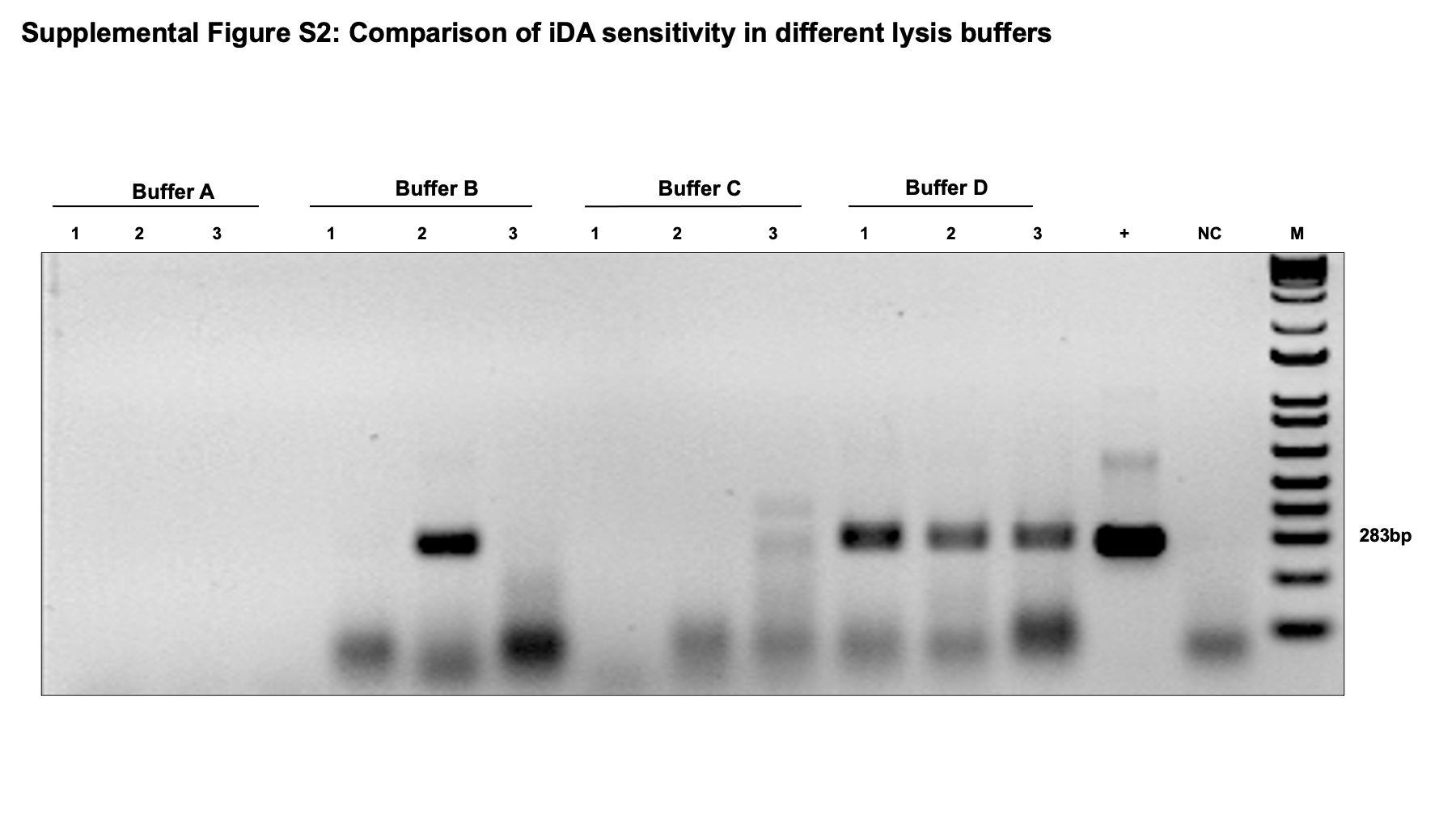

### SuppSlide3.tiff

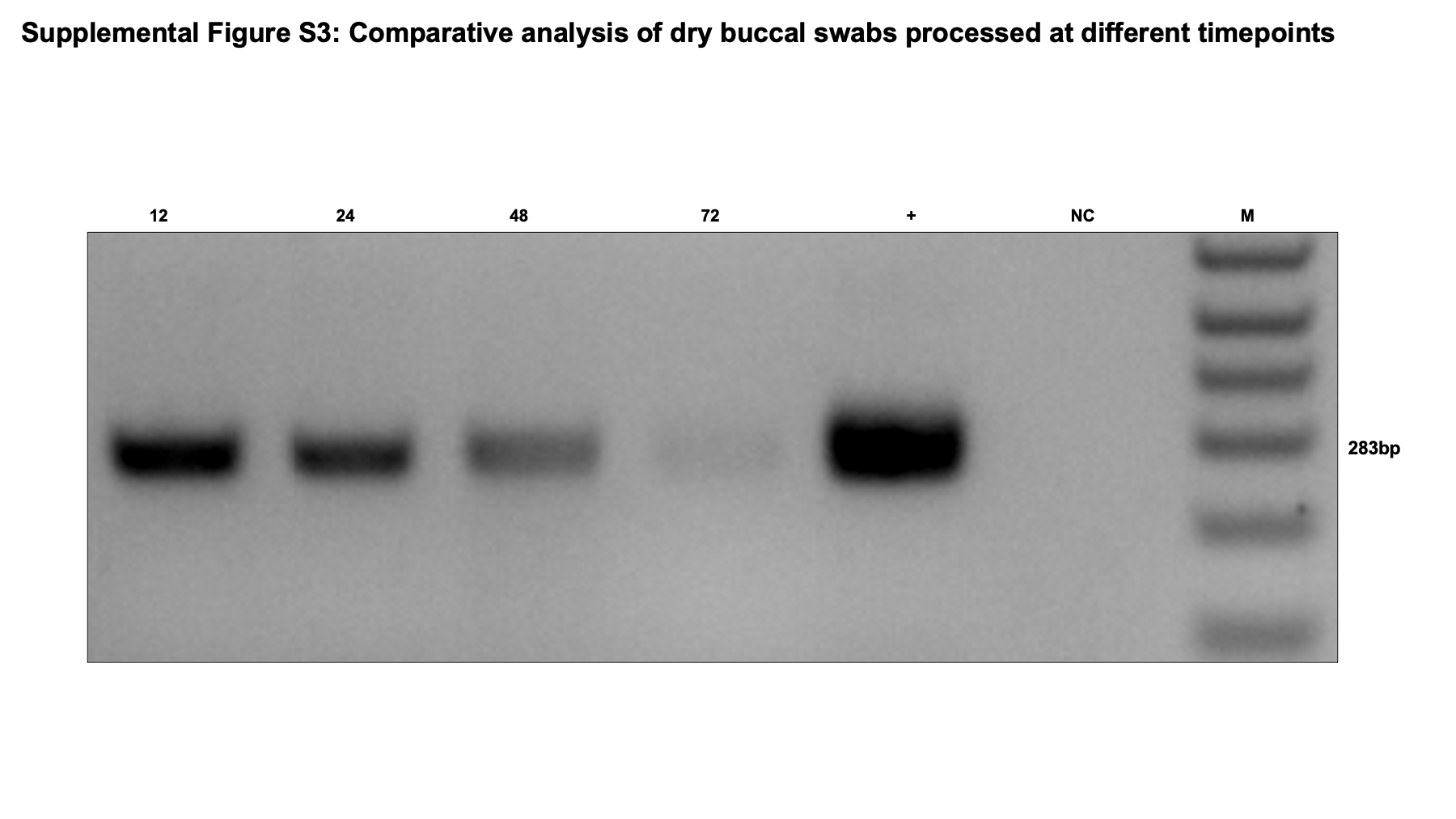

### SuppSlide4.tiff

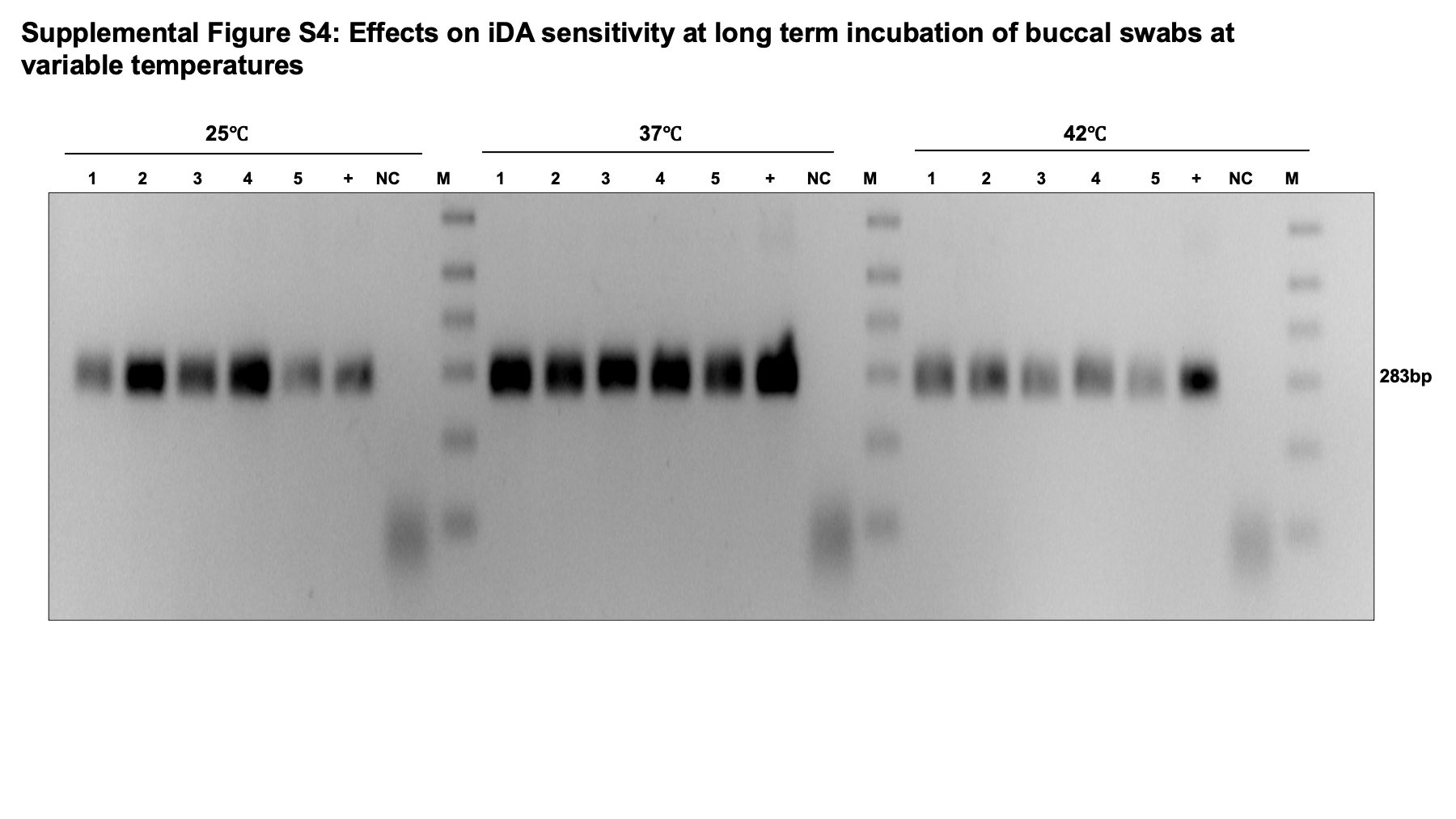
